# “Public attitudes to sharing government data with private industry: a systematic scoping review”

**DOI:** 10.1101/2022.06.07.22276084

**Authors:** Jackie Street, Stacy Carter, Belinda Fabrianesi, Rebecca Bosward, Lucy Carolan, Annette Braunack-Mayer

## Abstract

**Background:** Linking and analysing large volumes of health data in electronic records, datasets, registries and biobanks can provide detailed insights into the health profiles of individuals, communities, regions and national populations. Good governance for data sharing can be challenging, particularly when private sector organisations are the recipients of public sector data. Many jurisdictions have responded by instituting new regulations and laws related to data protection, responding to calls to promote data sharing and manage associated risks. This review integrates and interprets five years of research on public and patient perspectives on sharing publicly-held health data with private industry for research and development. Our review question was: what are community attitudes towards the use of government administrative health data by private industry organisations for therapeutic development?

**Methods:** We developed two logic grids: first, used terms describing citizens or patients, big data, private health sector and views or perspectives, second, used terms describing big data, social licence and public interest. We conducted a systematic literature search using electronic databases; PubMed, Scopus, CINAHL, Web of Science. Searches were conducted using Google Advanced and Google Scholar to identify grey literature

**Results:** A total of 6,788 articles were screened based on title and abstract. Full text screening was conducted for 224 articles and a total of 33 publications were identified. Across all studies, support for sharing administrative health data with private sector organisations was low. Background knowledge and lack of trust that data use would lead to public benefit were the most important reasons for low level of support.

**Conclusion:** Policymakers, data custodians and companies will need to establish robust safeguards including secure storage, anonymisation, monitoring and independent oversight, and institute and implement sanctions on misuse, if they are to secure legitimacy to share publicly-held health data with private industry for research and development.

**Registration:** *none available*.

**‘Contributions to the literature’:**

- Research shows governance for data sharing is challenging, particularly when private sector organisations are the recipients of public sector data. Globally jurisdictions have responded by instituting new regulations and laws related to data protection, data sharing and the associated risks.
- Bringing together research on ‘public attitudes towards the use of linked government administrative data by private sector organisations for therapeutic development’, this systematic review found that public support for sharing was low.
- These findings highlight key areas for policymakers, data custodians and companies to address before they can legitimately share publicly-held health data with private industry for research and development.

## Introduction

Data are increasingly central to contemporary life and work, including in the health sector. Linking and analysing the very large volumes of health data in electronic health records, research datasets, registries and biobanks can provide detailed insights into the health profiles of individuals, communities, regions and national populations.

It is widely acknowledged that governance for data sharing is important. It is also challenging, particularly when private sector organisations are the recipients of public sector data. (1-3) Concerns about private sector use of public data can, in part, be traced to the profit-driven nature of private sector companies. (4) Private companies are accountable to their shareholders and, at least in countries such as Australia, are required to serve the interests of shareholders (5), potentially conflicting with legislative requirements and public expectations that public sector administrative data are used in the public interest. (6)

Many jurisdictions have responded to these challenges by instituting new regulations and laws related to data protection, responding both to calls to promote data sharing (7-14) and to manage the risks associated with sharing data. (1, 2) For example, the European Commission, in 2018, adopted the General Data Protection Regulation (GDPR) giving individuals control over their personal data and instituting large fines for privacy breaches. Other countries, including the US, (13) Canada (8) and Australia, (9) are adopting legislative changes which will allow greater data sharing with the private sector under supervision, (7-15) acknowledging both the important contribution that the private sector makes to research and development of new pharmaceuticals and medical devices, and also noting the challenge that sharing data with the private sector poses for public trust in government.

In light of such proposed changes, it is surprising that so little attention has been paid to public views about sharing administrative health data sets with the private sector. The evidence that is available is scattered across papers that focus primarily on data sharing with other government agencies, with researchers or with non-government organisations, rather than specifically with the private sector. (1) Therefore, this paper brings together, for the first time, qualitative and quantitative research published in the peer reviewed and grey literature on public attitudes towards the use of linked government administrative data by private sector organisations for therapeutic development. Our review question was:

What are community attitudes towards the use of government administrative health data by private industry organisations for therapeutic development?

*Please refer to Additional file 1 for the PRISMA 2020 checklist/ reporting guidelines*

## Methods

### Search strategy

In designing and conducting the scoping review, we drew on the work of Arksey and O’Malley (16) and Peters et al. (17) Since we were primarily concerned with the breadth of existing literature in the area, we did not exclude papers based on quality but used a Mixed Methods Appraisal Tool (18) to note study limitations.

We developed two logic grids (population, concept, context, outcomes) for the study. The first search used terms describing citizens or patients, big data, the private health sector and views or perspectives, with these terms and relevant synonyms included in the searches. The second search used terms describing Big Data, social licence and the public interest, with these terms and relevant synonyms included in the Boolean searches. (For full details see Additional file 2, **Supplementary** Tables S1 and S2). Depending on the database, documents were sourced within the time period “last five years” or January 1st 2014 to April 1st 2019. We excluded papers published prior to 2014 because literature on sharing public health data with the private sector was limited prior to 2014. (1) Earlier studies would not necessarily reflect current community views and judgements about the public interest. Studies published within the time period, but which reported data prior to 2014, were included.

We conducted a systematic literature search using four electronic databases PubMed, Scopus, CINAHL, and Web of Science. In addition, searches were conducted using Google Advanced and Google Scholar, in particular to identify relevant grey literature. These databases were selected for their coverage of research with respect to the use of data analytics in health and, in particular, research on community attitudes to data sharing in health. In Google Advanced and Google Scholar, searches were restricted to the first 1000 hits. Additional ‘pearled’ relevant articles were extracted from the reference lists of included papers. Peer-reviewed and unpublished articles, reports, books and book chapters describing empirical research were included. Editorials or opinion pieces were excluded. There were no limitations on geographical location but only English language articles were included.

For the database searches, we iteratively developed a search strategy based on the logic grid. Our final search strategy is shown in Supplementary Tables S1 and S2. (Additional file 2, **Supplementary** Tables S1-S4).

### Inclusion/exclusion criteria

We screened title and abstract and, in the case of reports, the contents page using the criteria described in Table1. Articles were screened based on title and abstract (by authors JS, BF & RB). Full text screening was conducted by two authors (JS & BF). Where there was disagreement between the reviewers, the final decision for inclusion was made through discussion in the research team. Reference lists of included papers were reviewed and further articles identified.

*TO BE INSERTED HERE

(Additional file 3; **Table 1**: Inclusion and exclusion criteria for selection of articles)

### Data extraction and analysis

A template for data extraction was used to provide a consistent approach to extraction and reporting of the findings. Two reviewers (BF & JS) extracted: title, author name, year of publication, location(s), aim(s), focus, public engaged, specific patient group, sample size, health technology, methodology, models (consent, data linkage, public interest), case studies, overarching results – access of private companies to public data, under what circumstances can public data be shared with private companies, consent, storage, definition of social /social contract, definition of public interest/public benefits, and bias/limitations which related to the research questions. One reviewer, JS, inductively coded, without *a priori* codes, the included articles using N-Vivo to extract descriptive themes and develop analytical themes. A second reviewer, BF, coded two papers and used the extracted data and the research question to cross-check the coding and extracted themes. Differences were discussed and resolved.

**Table 2:**
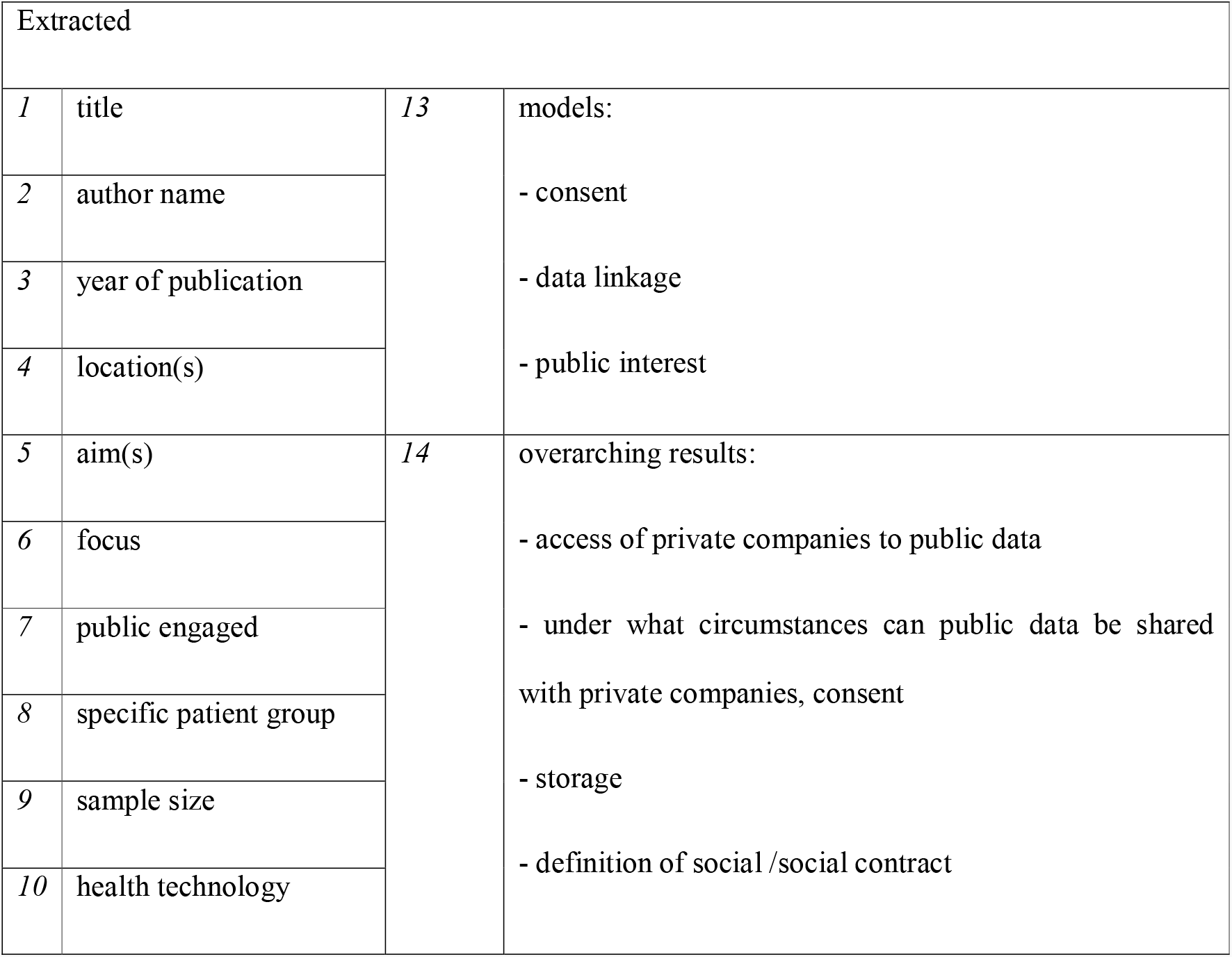
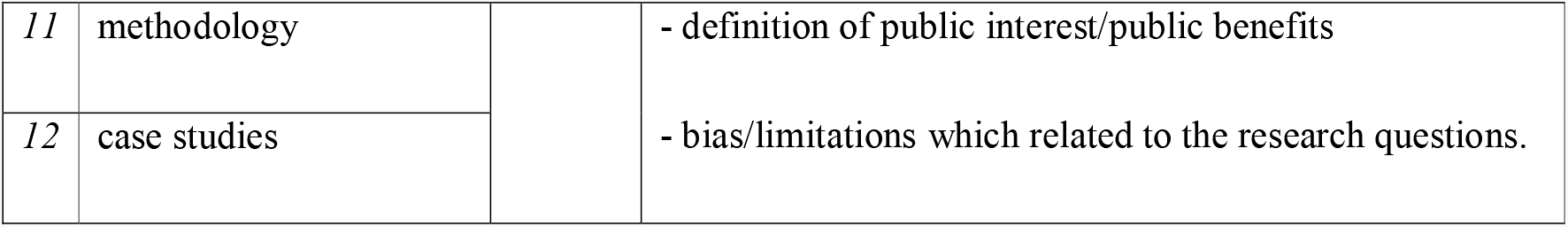
Extracted Terms

### Quality assessment

Study quality was assessed using the Mixed Methods Appraisal Tool (MMAT). This tool permits appraisal of the studies in mixed methods systematic and scoping reviews. It is designed to appraise the quality of the underpinning rationale and methodology. All studies meeting the inclusion criteria were assessed using seven criteria. The first two criteria established the existence of a clear research question while the second assessed the ability of the data collected to answer the question. The remaining five criteria were related to the study type e.g. qualitative/quantitative. Studies were given a score out of seven depending on how many of the criteria were met (18). Limitations of the studies are discussed.

## Results

A systematic literature search of the four electronic databases, Google Scholar and Google Advanced generated the following number of articles: PubMed (797), Scopus (1,768), CINAHL (389), and Web of Science (1,844), Google Scholar (1,990) and Google Advanced (293).

A total of 6,788 articles were screened based on title and abstract. Full text screening was conducted by two authors (JS & BF) for 224 articles (which included an additional seven pearled articles) and a total of 35 publications were identified. The flow chart in Figure 1 (see Additional file 4; Figure 1 PRISMA Flowchart) summarises the review selection process. Six papers scored below 5 on the MMAT assessment tool with only one scoring less than 3.

Of the 35 publications included in the review, 22 were peer-reviewed papers, 10 reports, two conference proceedings and one conference paper. Most papers reported on research conducted in the United Kingdom (n=17) and United States (n=6), with smaller numbers in New Zealand (n=3), Canada (n=2), international (n=3), Europe (n=2), South Korea (n=1) and Thailand (n=1). There were no studies identified from Australia, South America, Central America, Africa, Middle East, Russia or for much of Europe or Asia. Data collection within the included studies occurred during the period 2007-2018. A small number of studies did not report their data collection period. Participants included the broader public, affected patient groups, clinical stakeholders and private sector agencies. In this paper, we only report the views of patients, consumers and broader publics.

Fourteen of the studies were qualitative or deliberative, using focus groups, citizen juries, workshops, social assembly and interviews. Sixteen of the studies were quantitative using online and in person surveys, and there were three mixed method studies. Two research studies were reported in duplicate reports, the Data Futures study in New Zealand (19, 20) and the Life and Times Survey in Northern Ireland (21, 22). All but one of the studies (23) scored more than 3 in the MMAT quality assessment tool. Scores are shown in Table 3.

*TO BE INSERTED HERE

(Additional file 5; **Table 3**: A summary of the publications included)

### Public support for sharing administrative health data

In general, across all studies, support for sharing administrative health data with private sector organisations was low. Eight quantitative studies specifically asked about sharing health data with private sector organisations (Table 4). Most asked participants to rate how willing they were to share information with a range of individuals and groups; participants were generally less willing to share their non-identified data with private companies than with any other groups. (Table 4) The level of support varied widely, with lower levels of support for sharing identifiable data (15%) (52) and higher levels for a scenario involving the use of deidentified data to develop a cure for Alzheimer’s Disease (75%). (21) Similarly, in relevant qualitative studies, participants expressed less support for sharing personal health data with for-profit private sector organisations when compared with other groups involved in health research such as health care professionals, university researchers and non-profit organisations. (20, 25, 26, 33, 35, 42, 43)

There were two underlying reasons for a low level of support for data sharing with the private sector: background knowledge, and lack of trust that data use would lead to public benefit. With respect to background knowledge, participants were reportedly often surprised about the extent of data use and the possibility that their data might be shared with private companies. Because participants were not able to draw on existing understandings about drug and device development, and data use and sharing, the studies reported here either assumed familiarity with these issues or needed to include explanations of them (e.g. private and public health sectors, big data, data collection, wnership and sharing, health research governance and data safeguards). (4, 24, 27, 37, 38, 41-43, 47) Studies often reported comments such as:

> *Is this actually happening today, where they’re collecting a lot of data?* General Public, Focus Group 2, Toronto 2017 (43, p.E43)
>
> *Private companies have no need to have my medical information*. General Public, Glasgow. 2016 (4 p.36)
>
> *It says so they can predict what will make you ill or better. How? Are they god? How can they work all that out?* General public, Glasgow, 2016 (4, p.34)

In general, those studies that provided participants with opportunities to learn about data uses and safeguards reported that participants’ willingness to share with private companies increased when their knowledge increased. (4, 26, 42, 49) For example, in deliberative workshops in Scotland, although participants were wary of private industry involvement, they acknowledged their involvement as “a necessary evil”. (26, p.5)

The second background reason that participants rejected data sharing was a lack of trust that data use would lead to public benefit. (20, 24, 26) Some participants suggested data use can be “a blunt instrument” which may do more harm than good. (20, p.12) Distrust extended to government and universities but was highest for private industry. (20, 21, 24, 26, 42, 43, 47, 51) This lack of trust was not a simple relationship but rather was based on a complex web of experiences, beliefs and knowledge described in more detail below.

*TO BE INSERTED HERE

(Additional file 6; **Table 4**: Proportion of participants willing to share administrative health data with specific groups - all quantitative studies asking this question).

### Concerns about sharing data with private industry

The participants in the included studies were generally concerned about sharing their government health data. Some concerns were extensions of fears they had about sharing their health data with anyone, although their disquiet was intensified if the data sharing involved private industry. There were three main groups of concerns: about privacy, risk of harms, and use of public and personal data for private profits and interests. We discuss each of these in turn; more detail and sample quotes are presented in Supplementary Table S3. (Additional file 4, **Supplementary** Tables S1-S4)

### Privacy and confidentiality

Many participants were concerned about loss of privacy and breaches of confidentiality. (4, 20, 24, 27, 33, 35-38, 42, 43, 48) In some studies, the terms ‘privacy’ and ‘confidentiality’ were used interchangeably when describing participants’ views about these matters (1, 38, 48), whilst in other studies only ‘privacy’ or ‘confidentiality’ was used. (27, 33, 36, 43) One paper specifically noted that study participants used the terms interchangeably, although the authors chose to use the term confidentiality because it was the issue they were evaluating in their study (33). This paper suggested that participants viewed maintaining confidentiality as an expression of respect “between two parties exchanging information” and as a form of control, “a way of limiting how data about oneself may be used by others”, (33, p.965) with the former reflecting the duty of confidentiality and the latter the maintenance of privacy.

Across studies, loss of privacy and breaches in confidentiality were often related back to inadequate data security. Numerous studies reported concerns about data security, including the potential for leaks and hacking, particularly when sharing data with private industry. (21, 23, 36, 37, 41-43, 47-49, 52) Concerns were in some cases related to the security of electronically stored data generally (4, 20, 42), but also to the belief that both public and private systems were leaky or unstable. (33, 36, 48) This was based on knowledge of media reports of data breaches, (48) prior personal experience of such breaches, perceptions that hospital record keeping was disorganised, (42) and concerns about the stability of private companies. (36) There were also concerns about potential confidentiality breaches and loss of privacy through on-selling data to other companies, particularly for marketing purposes. (4, 24, 36, 41, 43, 49). The Wellcome Trust report on public attitudes to commercial access indicated that, in their study, participants believed “that no amount of security could ever totally remove the risks involved in sharing data”. (4, p.11)

### Risk of Harm

Concerns about data security were closely related to concerns about data misuse, expressed both in terms of general uneasiness about the use of participants’ data for purposes of which they were unaware and might oppose (48) and, more particularly, the considerable potential for harms to individuals through the use of identifiable health data. The latter included the use of their own data to disadvantage them in employment, insurance cover, and financial and health care services. (4, 20, 27, 33-36, 38, 41-43, 47) Participants were also concerned that even aggregate data might be used to stigmatise individuals based on their ethnicity or to segment, exploit or disadvantage vulnerable groups. (4, 27) Some participants saw sharing data as one more sign that “we are heading for a dystopian, surveillance-based society”. (4, p.60) In a study involving older Swiss adults, participants expressed concern about the potential use of data in eugenics:

> *So, my only concern is, it has once been talked about, that it could be used to create the perfect human*… *or*… *that everyone would have blue eyes or a standard type or for military purposes. Of course, that is a big topic. I would be absolutely against that*. Female, age 69, Switzerland (35, p.8)

Participants in this study were also concerned about becoming a “transparent citizen” with personal knowledge about them accessible and available to many and that thereby “people or institutions … might gain unpredictable powers by it”. (35, p.7).

### Use of data for private profit and private interests

Although many participants acknowledged that there could be a role for commercial organisations in therapeutic development, at the same time they were concerned about the interests held by private industries [23, 29, 38, 45, 55], their lack of accountability (20, 42) and their focus on profit-making. (4, 20, 26, 38, 42, 48) Across studies, the use of publicly held health data for private profit was a common concern for a range of reasons. Some participants simply disliked the idea that private companies were making profit from their data, particularly without some measure of reciprocity:

> *Business involved changes things a lot for me – I’m unhappy with businesses getting personal data as they profit but don’t have to give anything back*. (20, p.13)

Others believed that, unlike government organisations, private industry put their own interests ahead of the public good: (20, 26, 35, 42, 52)

> *Unfortunately, my belief is that when people start making a profit out of it that’s when the ethics start getting a little bit less and a little bit less as the profit margin goes up the less ethical you are the more money you earn*. Participant 1, person with diabetes, London, UK:2016 (52, p.85)

They were concerned that the private sector had less oversight and were less accountable to the public:

> *I’m fine with all of these organisations except businesses. Government usage is safer because there is responsible governance, but there is no corresponding obligation for private businesses who want to make a profit*. General public, New Zealand, including First Nations peoples, 2017 (20, p.13)

Some participants in Scottish deliberative workshops expressed a belief that private companies had suppressed past ‘cancer cures’ and would suppress results in future research to increase their profits. (26) In several UK studies participants expressed concern that pharmaceutical companies would have access to publicly held health data to develop new drugs which they could then sell back to the National Health Service (24, 41, 42) or on to others for profit. (4, 24, 38, 48, 49)

Participants in many studies differentiated between private companies using government health data under regulated conditions for public benefit and unfettered access for generation of profit. (4, 21, 24-27, 31, 35, 41-43) One parent of a child with a rare disease commented:

> *Big pharma…Are they doing it with my consent, looking at a group to identify, make progress, come up with treatments, understand conditions more – I’d be comfortable with that. Or are they just given free rein on my daughter’s medical records so they can stabilize business, play entrepreneurs, gamble on it – no that not OK*. Parent of patient, Sheffield, UK, 2016 (4, p.57)

Finally, some participants saw the issue as ownership of the data and that industry use of government data for private profit was an inappropriate use of their personal property (48)

#### Circumstances under which government health data may be shared

The concerns held by participants fed into their discussions about the conditions under which it would be acceptable for governments to share government-held administrative health data. Participants’ concerns about the purpose for data sharing led to a requirement that research should produce public benefits; concerns about data security led to requirements for secure data storage and de-identification; and concerns about misuse led to requirements which would restrict access and protect the data.

There was substantial agreement across studies about the circumstances under which government health data could be shared. The primary requirements were that:

- the research should produce public benefits;
- the data should be secure as possible, particularly that it be de-identified and securely stored;
- governance structures be in place to monitor and regulate access to data; and
- that due reciprocity should be considered.

There were a range of views about whether informed consent for data use was necessary and the form that consent should take. Here we discuss the principal conditions which participants believed were necessary before data should be shared. A full list of can be found online in Supplementary Table S4.

(Additional file 4, **Supplementary** Tables S1-S4).

### Public benefit

Although participants across studies considered it essential that the public should benefit from the sharing of public administrative data with private industry, what constituted benefit varied widely. (Table 5) In many studies (4, 20, 24-28, 31, 33, 41, 42, 51) public benefit was taken to mean something “would (at least probably) ultimately lead to benefits for healthcare” (24, p.716) including “finding cures for diseases and making new drugs available” (26, p.7) and monitoring the safety of new health technologies. Cancer, dementia and mental health and research that improved health and quality of life through preventive measures (see Table 5) were highlighted as important areas. Some participants questioned the need for private industry to be involved: e.g. in a UK study using deliberative workshops, participants valued monitoring for drug safety, but asked why the NHS could not conduct this work itself. (4, p.51)

**Table 5:** Conceptualisation of benefits through data linkage and use

| Concept | References |
| --- | --- |
| Disease diagnoses, treatments and cures (including specifically cancer, dementia, mental health and rare diseases) to benefit both individuals and groups | (4, 20, 21, 26, 28, 35, 37, 40-45, 47-53) |
| Improved population health and wellbeing including through prevention | (26, 35, 38, 41, 42, 48, 49, 52) |
| Monitoring the long-term safety and efficacy of drugs and treatments | (4, 37, 40, 41, 43, 48, 49) |
| Improving health services and health outcomes for vulnerable or disadvantaged populations | (4, 20, 24, 26, 37, 38, 52) |
| Creation and dissemination of new knowledge | [29, 41, 51, 52] |
| Improved allocation of resources, cost-effective care | (26, 37, 41, 49) |
| Promoting social good through contributing to research | (26, 41, 45) |
| Improved health policy | (20, 37) |
| Giving “children the best start in life” | (4, 26, 35) |
| Improving the lives of older people | (26) |
| Improving the natural environment | (26) |
| Benefits for the environment | (26) |
| Access to a wider skill set if private industry is involved | (43) |
| Ability to detect rare health events | (37) |

Participants with a history of personal or family illness were more likely to see data sharing, including with private industry, as essential to support development of new treatments. (28, 33, 50)

> *Patients are key to advancing research by providing data to researchers* - *the more information collected, the more it will promote advancement of research* - *in a rare disease like this, maximum participation is required for effective research*. Patients/relatives of patients with leukodystrophies. (28, p.7)

But, even here, there were differences of opinion. One participant expressed strong opposition to sharing with private industry stating: “The Pharma industry orients research in their own interests, not in the interests of patients.” (28, p.340)

Health benefit was also conceptualised as improving options for those with greatest need. For example, in deliberative workshops, Scottish participants acknowledged the merit in health research targeting “vulnerable groups who would potentially benefit the most” (26, p.7) and that benefits for small numbers of people could “lead to wider benefits for society”. (26, p.7)

### Regulation and safeguards

The participants in these studies wanted to see their data anonymised. Many studies did not discuss the potential for reidentification and it may be that many participants were unaware that there was potential for this to occur and, therefore, that they could never be completely anonymous. (20, 21, 23-25, 27, 30, 36, 37, 44, 47, 50, 52) From the participants’ perspective, anonymisation of government health data through removing personal information or aggregation was either a pre-requisite for acceptance of data sharing (4, 25, 27, 33, 36, 44) or greatly increased willingness to share. (20, 24, 35, 37, 38, 41, 43, 47, 52) For example, in a large UK survey (n=2017) anonymity was seen as the second most important condition for sharing health data with commercial organisations. (4) In a smaller study, although just under half the participants (patients with diabetes n=404) indicated they would be willing to share their government health data with pharmaceutical companies provided it was anonymised, this fell to 15% if identifiers were retained. (52) In several UK studies, anonymisation of data was seen as particularly important if data was to be shared with private companies: participants were concerned that identifiable information could be misused and it may still be possible to re-identify anonymised data. (4, 33, 48)

Most studies reported support for rigorous governance structures to monitor and regulate access to government health data. In many studies participants wanted multiple safeguards to be instituted (4, 20, 24, 28, 36, 38) and some participants suggested that private entities might gain trust if they were willing to “subject themselves to regulatory scrutiny”. (4, p.56). In particular, participants called for strict rules prohibiting passing data to third parties, anonymisation for data sharing, sanctions for misuse of data, secure data storage and oversight by an ethics committee. (4) Monitoring of individual access to the data by logging contact episodes was also suggested, the rationale being that this would serve as a deterrent for malpractice.

> *That would make me feel a bit more comfortable because they would know, if for any reason the system had been abused, not that it would be but they would know…There’ll be a shortlist of people who have accessed, it would be a deterrent of abus*e. General Public, Belfast, UK (4, p.63)

Secure storage (4, 20, 21, 33, 36, 43, 48, 49, 52) and independent oversight. (4, 24, 25, 36-38, 43, 48, 49) were widely recognised as essential conditions for data sharing. One participant in a European study likened the necessary controls to those found in the banking sector:

> *I’m just trying to say there is this framework, you know we say that there is a governance system in place which will protect the patient and we can look at them like we do the financial institutions and we’re quite happy with how they exist, well they’re quite well developed. There’s a framework around this and we want some assurance*. Patients/parents of patients with a rare disease, Europe, (36, p.1,045)

One individual writing in response to attempts to share public administrative data sets in the UK said:

> *I want the data to be supervised by an independent forum of individuals whose remit is to follow strict published ethical guidelines relating to sharing, selling and profit making by the use of my data*. UK, Comment posted to website Care.data, 22.01.14 (48, p.184)

Scottish focus group participants suggested members of the public should have a role in an oversight committee “ensuring accountability and protecting public interests”. (24, p.720) Clear explanations about how data would be recorded, anonymised and stored was seen to be helpful in building support for data sharing:

> *He explained to me that basically there’s only one location where there’s a cross-reference between the name of the participant and the identification process they’re using on each individual patient’s, or study participant’s, file. So I don’t have any issues with tha*t. ClinSeq#120, NIH genomic research registry participant, USA (33, p.967)

Channelling information through trusted entities may increase public acceptance of data sharing but some participants indicated that it could also erode trust in health care providers.

> *I do not trust the government with my data, and now I cannot trust my doctor o[r] the wider NHS*. Comment posted to Care.data website, 05.05.14, (48, p.183)

### Reciprocity

Across studies, many participants called for a level of reciprocity from private companies receiving administrative health data. This included that there be openness and transparency about how data were stored, shared and used (4, 24, 27, 28, 33, 35, 38) as well as public release of the research findings. (4, 21, 24, 28, 30, 33, 38, 47) In addition, some participants called for access for research contributors to their own data (28, 52) and for patient participants to any new technologies resulting from the research. (41) Some believed that private industry should pay for data access or, where applicable, deliver some of the ensuing profit back into the public system. (21, 22, 38, 41-43)

### Consent

Consent was an important consideration in many studies but the need for and type of consent was highly contentious. (4, 20, 27-29, 31, 33, 36) Some participants were comfortable with government health data sharing with no consent - particularly where data were anonymised or aggregated and the research of high public benefit - whereas others wanted explicit consent on every occasion. The majority of participants in two deliberative studies that explicitly focused on sharing with private industry started from a position of requiring informed consent but moved to no-consent required for cases with high public benefit where consent was impossible or prohibitively expensive. (4, 49)

Participants in one of the studies called for better communication around data use without consent (4) and some participants suggested that their ongoing reservations related to a lack of clarity about the personal implications of sharing their data with private industry. (4, 33) Some members of the NICE Citizens’ Council became more concerned following deliberation. One participant reflecting this shift said:

> “I’ve completely changed my mind. When I first started [the meeting] I thought ‘yeah’, but now all I can think of is cons.” NICE Citizens Council (41, p.23)

## Discussion

This scoping review integrates and interprets five years of research on public and patient perspectives on sharing publicly-held health data with private industry for research and development. Many studies noted that publics and patients have limited opportunities to understand the nature and extent of data use, sharing and protection; researchers consequently provided information to participants as part of the research process in a number of the included studies. Our findings are consistent with other systematic reviews of international perspectives on health data sharing. (1, 54, 55) Our first conclusion is:

> *1) People support sharing public sector health data for health research in general, but are less willing to share with the private sector for research and development*.

This conclusion is consistent with research on general public attitudes to sharing, and with research on public attitudes to: sharing genomic data (56)(56), secondary use of clinical trial or public health study data (57), and “broad consent” for data sharing. (2) We thus suggest this central finding is robust, at least within the UK and North American settings (where most research has been conducted). (1, 54, 55) Further research is necessary to understand generalisability to other contexts. Participants expressed concerns regarding perceived *vulnerabilities in data security*, potential for *misuse of data*, and potential for *discrimination* against, or *stigmatisation* of, individuals or groups.

Quantified measures of support for sharing health data with private actors ranged from 16% to 75% (i.e. from on-balance rejection to on-balance acceptance). This leads to our second conclusion:

> *2) Public support for sharing public sector health data with the private sector is possible, but is more strongly conditional than support for sharing with public sector and non-profit actors*.

Our review suggests that any government, agency or company considering public-private health data sharing needs to develop a social licence, and that this will require providing, negotiating and assuring granular commitments regarding what data are shared, why they are shared, how identification is handled, and the governance and accountability mechanisms in place (Table 4). In particular, publics expect that data sharing should produce *public benefit*, that safeguards including *secure storage, anonymisation, monitoring and independent oversight* would be in place, and that *sanctions* would follow misuse. While other reviews have reported similar expectations with respect to sharing in general, (1, 54, 55) this review adds clear evidence that these *expectations become far stronger when data are shared with private industry*. These higher expectations for public-private sharing were informed by disquiet about using a public resource for private profit. A profit motive was seen to exacerbate the risks that participants were concerned about: security, misuse, and potential for harm (e.g. via stigmatisation or discrimination). (4, 49) For-profit use was also seen to undermine a non-negotiable and central condition for sharing: that data be used for public benefit or the public good (which could include aggregative benefit to individuals).

Similar to other reviews (1, 54, 55), we found public benefit was also highly relevant to consent requirements. This leads to our third conclusion:

> *3) Preferences regarding consent models reflect judgements regarding two underpinning considerations: whether the parties involved can be trusted, and the likely public benefit from sharing data*.

Generally higher trust and/or higher public benefit allowed acceptance of consent models with lower individual control. However, for some participants, if data sharing involved private industry, explicit consent was essential even if public benefit was high. In systematic reviews of public attitudes towards sharing of genomic data, and secondary use of research data, individual-level consent has been seen as a way to address concerns (e.g., about security, misuse and harm) (56, 57). Note, however, it is generally considered logistically infeasible to obtain individual consent for each act of sharing administrative health data. Also, individual-level consent may make a dataset less representative, or systematically biased, due to uneven non-participation, making findings untrustworthy. If active consent is seen as the *only* way to secure socially acceptable sharing with private industry, such sharing may be impossible. Further research is needed to explore alternative ways to develop the social licence to share data with the private sector. For example, our own research using deliberative methods suggests that informed citizens may be willing to accept sharing their data, including with private industry, provided sharing is tightly regulated and in the public interest (58).

## Study Limitations

Six of the articles scored less than 5 using the MMAT tool and 3 had a score of less than 4. One paper, reporting a single focus group, had a score of 1 and, although noted, was not included in the findings of the review.

## Conclusion

Overall, this review has indicated key areas for policymakers, data custodians and companies to address if they are to secure legitimacy for the sharing of publicly-held health data with private industry for research and development. Each program of data sharing would require carefully negotiated commitments to establish safeguards including secure storage, anonymisation, monitoring and independent oversight, and to institute and implement sanctions on misuse. Patients and publics would require assurance that data use will lead to public benefit, and that all actors and systems involved are trustworthy. These things may be achievable, but two problems currently appear intractable. When data are shared for private profit, publics—quite reasonably—recognise a conflict of interest and are thus less likely to trust safeguards or assurances. Without both trust and assurance of public benefit, public-private data sharing is unlikely to be supported. In situations of lower trust and/or lower public benefit, publics are more likely to expect that they will have an opportunity to provide individual consent for instances of sharing, which is potentially fatal to the data sharing enterprise. More research is required to determine how this conundrum can be addressed.

## Supporting information

Cover letter

PRISMA reporting guidelines

Table 1 Inclusion _Exclusion Criteria

PRISMA Flow Chart

Table 3 Included articles

Table 4 Participants willing to share

## Data Availability

All relevant data are within the manuscript and its Supporting Information files.

## Declarations

### Ethics approval and consent to participate

Not applicable

### Consent for publication

Not applicable

### Availability of data and materials

Data sharing is not applicable to this article as no datasets were generated or analysed during the current study.

### Authors’ contributions

JS conceptualised the study, contributed to data collection, analysis and interpretation, drafted and approved the final manuscript.

SC contributed to the study concept and design, data analysis and interpretation, drafting the manuscript and approved the final manuscript

BF contributed to the study concept and design, data collection, data analysis and interpretation, drafting the manuscript and approved the final manuscript

RB contributed to data collection, data analysis and interpretation, drafting the manuscript and approved the final manuscript

LC contributed to the study concept and design, data analysis and interpretation, drafting the manuscript and approved the final manuscript

ABM conceptualised the study, contributed to data analysis and interpretation, drafting the manuscript and approved the final manuscript. ABM is responsible for obtaining the funding for the project and is the corresponding author.

## Acknowledgements

The authors would like to thank the Population Health Research Network for funding the project, The content is solely the responsibility of the authors and does not represent the views of the Population Health Research Network.

