## Supplementary tables S1-S4 for "“Public attitudes to sharing government data with private industry: a systematic scoping review”"

### Supplementary Table S1: Logic grid

**Logic grid – Research question one**

The logic grid below was developed to explore the first research question:

What are community attitudes towards the use of government health data by private sector organisations for therapeutic development?

| Population | Concept or phenomena of interest | Context | Outcome/themes |
| --- | --- | --- | --- |
| Community  (communit* OR patient* OR public OR citizen* OR client* OR consumer*) | Big data  (“health data” OR “health information” OR “Big data” OR “information sharing” OR “Data mining” OR “Data analytics” OR “Data linkage” OR “data sharing” OR “electronic health record” OR “electronic health data” OR “electronic medical record” OR “electronic medical data” OR “electronic patient record”) | Therapeutic development (medicine*OR “health technolog*” OR device* OR therapeutic*) | Attitudes, views or perspectives  (attitude* OR perspective* OR view* OR opinion*) |
| Community  (communit* OR patient* OR public OR citizen* OR client* OR consumer*) | Big data  (“health data” OR “health information” OR “Big data” OR “information sharing” OR “Data mining” OR “Data analytics” OR “Data linkage” OR “data sharing” OR “electronic health record” OR “electronic health data” OR “electronic medical record” OR “electronic medical data” OR “electronic patient record”) | Private sector  (“private sector” OR industry OR commercial) | Attitudes, views or perspectives  (attitude* OR perspective* OR view* OR opinion*) |

**Logic grid – Research question two**

The logic grid below was developed to explore the second research question:

What is the public interest and social licence for the use of government health data by private sector organisations for therapeutic development?

| Concept/ phenomena of interest | Outcome/themes |
| --- | --- |
| Big data  (“health data” OR “health information” OR “Big data” OR “Data mining” OR “Data analytics” OR “Data linkage” OR “data sharing” OR “information sharing” OR "electronic health record" OR "electronic health data" OR "electronic medical record" OR "electronic medical data" OR "electronic patient record") | Social licence and public interest  (“social licence” OR “public interest” OR “public good” OR “public benefit” OR “social trust” OR “social value” |

### Supplementary Table S2: Database search strategy

Time period: Last five years OR January 1st 2014 to April 1st 2019

Language: English only

Geographical location: All

Article type: journal article, report, book, book chapters

| **Database** | **Search** | **Articles (n)** |
| --- | --- | --- |
| SCOPUS | ( TITLE-ABS-KEY ( "health data" OR "health information" OR "big data" OR "data analytics" OR "data linkage" OR "data sharing" OR "data mining" OR “information sharing”) ) AND ( TITLE-ABS-KEY ( attitude* OR perspective* OR view* ) ) AND ( TITLE-ABS-KEY ( medicine* OR "health technolog*" OR device* OR therapeutic*) ) AND ( TITLE-ABS-KEY ( public OR communit* OR consumer* OR patient* OR client* ) )  ( TITLE-ABS-KEY (“electronic health record” OR “electronic health data” OR “electronic medical record” OR “electronic medical data” OR “electronic patient record”) ) AND ( TITLE-ABS-KEY ( attitude* OR perspective* OR view* OR opinion*) ) AND ( TITLE-ABS-KEY ( medicine* OR "health technolog*" OR device* OR therapeutic*) ) AND ( TITLE-ABS-KEY ( public OR communit* OR consumer* OR patient* OR client* ) )  ( ( TITLE-ABS-KEY ( "health data" OR "health information" OR "big data" OR "data analytics" OR "data linkage" OR "data sharing" OR "data mining" OR "information sharing" ) ) OR ( TITLE-ABS-KEY ( "electronic health record" OR "electronic health data" OR "electronic medical record" OR "electronic medical data" OR "electronic patient record" ) ) ) AND ( TITLE-ABS-KEY ( "social license" OR "public benefit" OR "public good" OR "social trust" OR "social value" ) ) AND ( TITLE-ABS-KEY ( health OR medic* OR therapeutic OR clinical ) ) | 995  703  70 |
| Medline (PubMed) | ((("Big Data"[Mesh] OR "Data Mining"[Mesh] OR "Data Science"[Mesh] OR "Information Dissemination"[Mesh] OR "health data"[TW] OR "information sharing"[TW] OR "health information"[TW]) AND ("Health Knowledge, Attitudes, Practice"[Mesh] OR "Attitude to Health"[Mesh] OR view[TW] OR perspective[TW] OR attitude[TW] OR opinion[TW])) AND (drug[TW] OR medicine[TW] OR "health technology"[TW] OR "medical device"[TW] OR therapeutic[TW])) AND (community[TW] OR patient[TW] OR patients[TW] OR public[TW] OR citizen[TW] OR citizens[TW] OR client[TW] OR consumer[TW])  ((("electronic health record” OR “electronic health data” OR “electronic medical record” OR “electronic medical data” OR “electronic patient record”) AND ("Health Knowledge, Attitudes, Practice"[Mesh] OR "Attitude to Health"[Mesh] OR view[TW] OR perspective[TW] OR attitude[TW] OR opinion[TW])) AND (drug[TW] OR medicine[TW] OR "health technology"[TW] OR "medical device"[TW] OR therapeutic[TW])) AND (community[TW] OR patient[TW] OR patients[TW] OR public[TW] OR citizen[TW] OR citizens[TW] OR client[TW] OR consumer[TW])  (("Electronic Health Records"[Mesh] OR "Big Data"[Mesh] OR "Data Mining"[Mesh] OR "Data Science"[Mesh] OR "Information Dissemination"[Mesh] OR "health data"[TW] OR "information sharing"[TW] OR "health information"[TW])) AND (("social license" OR "public benefit" OR "public good" OR "social trust" OR "social value") | 525  201  71 |
| WOS | (TS=("health data" OR "health information" OR "big data" OR "data analytics" OR "data linkage" OR "data sharing" OR "data mining" OR "information sharing")) AND (TS= (attitude* OR perspective* OR view*)) AND (TS= (medicine* OR "health technolog*" OR device* OR therapeutic*)) AND (TS=(public* OR communit* OR consumer* OR patient* OR client* OR citizen*))  (TS=("electronic health record” OR “electronic health data” OR “electronic medical record” OR “electronic medical data” OR “electronic patient record”)) AND (TS= (attitude* OR perspective* OR view* OR opinion*)) AND (TS= (medicine* OR "health technolog*" OR device* OR therapeutic*)) AND (TS=(public* OR communit* OR consumer* OR patient* OR client* OR citizen*))  TOPIC = ("health data" OR "health information" OR "big data" OR "data analytics" OR "data linkage" OR "data sharing" OR "data mining" OR "information sharing" OR "electronic health record” OR “electronic health data” OR “electronic medical record” OR “electronic medical data” OR “electronic patient record”) AND TOPIC = ("social license" OR "public benefit" OR "public good" OR "social trust" OR "social value") | 1607  138  99 |
| Cinahl | ("health data" OR "health information" OR "big data" OR "data analytics" OR "data linkage" OR "data sharing" OR "data mining" OR "information sharing") AND (attitude* OR perspective* OR view*) AND (medicine* OR "health technolog*" OR device* OR therapeutic) AND (public* OR communit* OR consumer* OR patient* OR client* OR citizen*)  ("electronic health record” OR “electronic health data” OR “electronic medical record” OR “electronic medical data” OR “electronic patient record”) AND (attitude* OR perspective* OR view* OR opinion*) AND (medicine* OR “health technolog*” OR device* OR therapeutic) AND (public* OR communit* OR consumer* OR patient* OR client* OR citizen*)  ("health data" OR "health information" OR "big data" OR "data analytics" OR "data linkage" OR "data sharing" OR "data mining" OR "information sharing" OR "electronic health record” OR “electronic health data” OR “electronic medical record” OR “electronic medical data” OR “electronic patient record”) AND ( "social license" OR "public benefit" OR "public good" OR "social trust" OR "social value" ) | 295  70  24 |
| Google Scholar | ("patient data" OR "data analytics" OR "big data" OR "data sharing" OR “electronic health record”) AND (community OR consumer OR public OR patient) AND (attitude OR view OR perspective OR opinion) AND health  health AND ("public interest" OR "social license") AND ("health data" OR "electronic health record" OR "data analytics" OR "big data" OR "data linkage" OR "data sharing") AND (private OR commercial OR industry) | First 1000  First 1000 |
| Google Advanced | (community OR consumer OR client OR public) AND ("health data" OR "data analytics" OR "information sharing" OR "big data" OR "data mining" or "data linkage" OR "data sharing") AND (private OR commercial)  health AND ("public interest" OR "social license") AND ("health data" OR "electronic health record" OR "data analytics" OR "big data" OR "data linkage" OR "data sharing") AND (private OR commercial OR industry) | 206  123 |

### Supplementary Table S3: Summary of concerns about data sharing with sample quotes

| Concern | Sample quote from a participant or from text in the article |
| --- | --- |
| Data security, data leaks and hacking | Best practice guidelines are ﬁne – but people are just fallible. One person, without malice, discloses 1,000 people’s sensitive data through an insecure email. Mistakes just happen, but once it’s out there, you can’t get it back. General Public, workshops, New Zealand. (19, p.13) |
| Loss of privacy lack of respect for confidentiality and privacy | ‘Drunk driving’s still an offence even if you don’t have an accident, because society thinks the potential of harm is sufﬁcient. Are you seriously saying that medical conﬁdentiality only matters in retrospect if the failure causes explicit harm, rather the potential for harm?’ Letter to The Guardian, UK. 19.08.14. (31, p.182) |
| Data exposure causing stigmatisation | A small minority of participants described concern about risk to their privacy, speculating that patients with more sensitive health conditions may be "more guarded of what happens with their health information" (Participant #11) due to fear of stigmatization. Affected patient groups, UK. (32, p.5) |
| Surveillance | Some participants felt that we are heading for a dystopian, surveillance-based society. For them, whether the data is anonymised or not was almost not relevant because they were worried about the future risks to privacy rights as a result of so much increased data sharing of any kind. General public, UK. (25, p.11) |
| Use of data to discriminate in insurance | ‘They do penalise people with illnesses. [Like] car insurance. My daughter’s a prime example - she’s got MS but she has to have her driving licence renewed every three years and she pays a higher premium. She’s got MS but she is just as capable of driving as anyone’. – Participant, general public, Swansea, UK. (5, p.32) |
| Use of data to discriminate by an employer | ‘Yaa, the people who suffer from a disease, they want... they fear that it will become public, I think it is connected to their job, for example if they have an increased risk for diabetes, they might not get the job, since the employer might be worried that they will get sick, I guess that plays a role’. Participant, female, age 71. (21, p.8) |
| Use of data for marketing | [Clinical trial participants] expressed concern about their identifiable information being used by third parties, such as telemarketers and insurance agencies, for purposes unrelated to health research. Affected patient groups, Focus groups, Thailand. (34, p.5) |
| Suppression of data for political ends | ‘You could say the same about the government – does the government really want to ﬁnd cures? Because, at the end of the day, the longer people live, the more it costs to give them everything: pensions, health service, and what have you. So, do they want us to be living to 100 years old? Probably not’. Participant, Male, Focus group 3, Aberdeen, Scotland. (22, p.6) |
| Use of data to promote the interests of academic researchers | M2. ‘It would depend where the university was getting the funding from. There’s a chance the university could be getting their funding from a pharmaceutical company. F2: Again, that’s trust, isn’t it? You’re trusting those researchers to be ethical with their ﬁndings….’. Participant, focusgGroup 3, Glasgow, Scotland. (22, p.5) |
| Use of data for purposes which are not in the public interest or for public benefit | ‘If there is such a partnership, I refuse to participate in the database. The pharma industry orients research in their own interests, not in the interests of patients’. Rare disease patient/family member, survey, Europe. (12, p.8) |
| Use of data for eugenics | ‘So, my only concern is, it has once been talked about, that it could be used to create the perfect human... or... that everyone would have blue eyes or a standard type or for military purposes. Of course, that is a big topic. I would be absolutely against that’. - Participant, female, age 69, interviews, older adults, Switzerland. (21, p.9) |
| Use of data to deny treatment | ‘And then they combine all that together, and they say, okay, well, this person has got this and this and this. Wasting medication or treatment or whatever on this person, beyond this age is useless. Let’s just let this person die’. - Participant, focus group 1, Thunder Bay, Canada. (18, p.E43) |
| Use of data to generate profit | Pharmaceutical companies hold a lot of power and the potential for life or death, and make huge profits out of life or death situations, one person suggested. Another said, We feel as a society we are at the mercy of pharmaceutical companies because they usually put profit before people. - General public, Citizens’ Council, UK. (31, p.25) |
| Lack of transparency in how data is used | ‘A deliberate decision was made to sell our data to a private company for purely commercial use, having nothing whatsoever to do with improving medical care or NHS services. This government has lied about its intentions for the NHS, and continues to lie and obstruct information about what it has done with it, and what is going on as we speak. This is all really too bad, because the existence of a large database like the NHS has such obvious beneﬁts. However this is only where the NHS remains a public service, and where the information is used to beneﬁt the public. NOT commercial businesses’. - Letter to The Guardian, UK, 18.08.14. (41, p.184) |
| Re-identification of anonymised data | ‘I am concerned that even anonymised information could be combined with other information that’s easily available to de-anonymise and identify me. I’m also concerned that other moves that are planned for the future will further erode patient conﬁdentiality beyond what has already been published’. - Comment posted to UK Care.data website. (41, p.182) |
| Concern that data may be sold on to others | ‘In particular, it seemed to go against natural justice that a company could repurpose data originally generated from the public, and make money again and again from the same dataset’. – Participant -general public, UK. (25, p.59) |
| Erosion of trust in health care professionals | ‘Patients will not conﬁde in their doctor, certain personal information, that may be absolutely necessary for diagnoses and treatment, knowing that outside agencies may have access to it’. - Comment posted to the UK Care.data website, 26.02.14. (41, p.183) |

### Supplementary Table S4: Summary of conditions which a proportion of participants believe needs to be met before data should be shared with private companies with sample quotes

| Condition (references) | Sample quote from a participant or from text in the article |
| --- | --- |
| Purpose |  |
| That there be clear public benefit (16, 17, 19, 21, 22, 24, 25, 30, 31, 33, 34, 39) | The involvement of a commercial organisation was seen as fairly easy to accept when participants could see clear potential for patients, society and future generations to benefit. This meant Linking data in the NHS and monitoring the safety of drugs were acceptable and seen as valuable. — General public, UK, 2015. (25, p.51) |
| That the use of the data be ‘ethical’ (20, 24) | ‘While I may be willing to share all my data for the purposes of improving health for the world at large, I ﬁnd the language used vague -most probably- on purpose. There is no guarantee that my data will be used ethically, All it says is that there are ‘‘strict rules to protect’’ privacy. I want strict rules to protect my data from being used in research relating to the creation, marketing or deployment of weapons; I want my data to be protected from being sold to or shared with companies which engage in the patenting of genome products; I want my data to be protected from being sold to or shared with companies that engage in abusive hiring practices here and abroad; I want my data to be protected from being sold or shared with companies or individuals that treat the environment with contempt; I want my data to be protected from being sold to or shared with companies that have unacceptable top executive salaries; and this is just a sample. I want the data to be supervised by an independent forum of individuals whose remit is to follow strict published ethical guidelines relating to sharing, selling and proﬁt making by the use of my data’. — Comment posted to UK Care.data website 22.01.14. (41, p.81) |
| That sharing of data not undermine equitable distribution of resources (22) | This related to wider discussions around the ways in which beneﬁts of health research are realised and a widely held perception that currently the beneﬁts are not realised equitably across society and that diﬀerent groups or people in diﬀerent locations across the UK experience health services diﬀerently as well as experiencing diﬀerent health outcomes. Throughout these discussions a recurring theme was that the potential beneﬁts of health research were not always or consistently realised. A range of factors were noted as limiting the realisation of public beneﬁts from health research, these included commercial interests, political priorities and limited public funding. — General public, workshops, Scotland, 2018. (22, p.6) |
| Avoiding Harm |  |
| That sharing the data causes no harm to individuals or society (25) | ‘Red lines: anything that risks personal harm, especially to vulnerable individuals’. - Participants describing circumstances under which data sharing should not occur, general public, survey, UK, 2016. (25, p.12) |
| Not for Profit |  |
| That public companies not overly profit from the data sharing at the expense of the public (12, 16-19, 21, 22, 31, 33, 50) | The majority of respondents (62 per cent) chose: “Any proﬁt made from research carried out using linked information should be invested into public services”. Only 8 per cent chose “Any proﬁt made from research carried out using linked information should be kept by those carrying out the research”. — General public, Scotland. (16, p.11)  ‘I guess I just think maybe they [the private sector] could fund their own research. I’m not sure the taxpayers should pay for it. But I guess, as you said, if they’re giving us an appropriate price or a better drug being released then I guess it’s okay’. - Participant, focus groups, Toronto, Canada, 2017. (18, E43) |
| That private companies pay for access to data (24, 31) | ‘I would like them to get access to patient records but they should pay a fee to get the data as they will make a profit out of any new drug’. Participant, General public, Survey, Northern Ireland. (24, p.20) |
| Consent |  |
| That consent be obtained before data is shared (12, 18, 19, 24, 25, 31, 32, 34, 38-42) | Council members suggested that systems should be established to ensure that researchers are transparent from the outset, with informed consent procedures requiring them to tell study participants how their data will be used and who might have access to it (including whether their data may be sold to other organisations), how long data will be kept and what will happen to data once the research study has finished. Explanations should be kept simple to ensure study participants can understand and they should be given written information. Informed consent procedures should include information about what a researcher plans to do with data if they discover incidental findings about a participant’s health and wellbeing. — General public, Citizens Council, UK, 2015. (31, p.27) |
| That participants have the option to opt out from data sharing (19, 21, 31) | ‘… it has to be voluntary, because we have to respect different personal inhibition thresholds in terms of sharing data… you cannot force someone or share data behind someone’s back. Out of respect’. - Participant, male, age 74, interviews Swiss older adults. (21, p.9)  Overall, several Citizens Council members considered it was important to allow individuals privacy and the right to choose to share their data because they considered this was important to society as a whole. They felt society was stronger if people had freedom of choice. - General public, Citizens Council, UK, 2015. (9, p.39) |
| That participants be well informed in the consent process (19, 20, 31, 33, 34, 38) | ‘We have to take time to explain until they (the clinical trial participants) understand. The bulls will not pull the cart and take offright after you connect the cart to them. As for us, we have been committee members for many years so once you explain, we understand immediately. New people will not understand it…Carefully explain one by one step by step.’ - CAB2, Community ethics advisory board member, male, focus groups, Mae Sot on the border of Thailand and Myanmar, Thailand, 2018. (34, p.6) |
| That consent procedures indicate what a researcher will do if they discover incidental findings relevant to a participant’s health and wellbeing. (31) | ‘Informed consent procedures should include information about what a researcher plans to do with data if they discover incidental findings about a participant’s health and wellbeing’. — Participant, general public, Citizens Council, UK, 2015. (31, p.27) |
| Reciprocity |  |
| That the public be informed about how and why the data is being shared and the potential known risks of sharing the data (19-21, 25, 33) | ‘Guess what? Maybe we, as a generation, need to go out on a limb a little bit here, but it’s that proper education of how this is going to be used, and proper thanks’. — Patients, interviews, USA. (33, p.544) |
| That participants in have access to their own data (29) | The majority of respondents said that they would prefer to have full access to their own medical history (91.52%) rather than limited access to their health information (4.09%). — Public and patients, survey, London, UK. (29, p.83) |
| That outcomes from any research involving their data be made public (12, 20, 24, 27, 32, 33) | ‘I just love this idea [Dynamic Consent], the updates they’re great. If I was involved with something [research] and it got published, I could go on the Internet and click on that [dynamic interface] and it would give me all the published papers on it’. — Participant #4, patients, interview, UK, 2016. (32, p.7) |
| That participants in research have access to the developed technology if it proves useful (31) | The group considering a new treatment for a rare condition were concerned that a patient might contribute to research by sharing their data but might no longer have access to or be able to afford the drug once it was approved and not available as part of a research study. — General public, Citizens’ Council, NICE, UK, 2015. (31, p.22) |
| That the benefit companies gain through data sharing be returned to the public in some way e.g. lower drug prices (17, 18) | ‘[If they] explain to me that the database is not only for medical purposes but would also get us access to more medical [services] in terms of the way the commissioning is taking place, then yes, you are making a good case to get me on the database, but if you are saying that, oh, I should just provide my [information] what’s this all this research going on for?’ — Participant, FG13, focus groups, patients, UK. (17, p.9) |
| Data Security and Confidentiality |  |
| That the data be securely stored (18-20, 24, 25, 29, 37, 38, 41) | ‘Well I think because it’s health data, it’s really important to keep it safeguarded. It’s not just some random information. It’s personal information. Really personal information’. — Group 2, focus groups, general public, Toronto, Canada, 2015. (18, p.E43) |
| That personal privacy and confidentiality be respected and maintained (12, 18, 20, 21, 27, 29, 31) | Council members considered there was no difference in the need to protect data and keep it confidential depending on who was collecting and using data. Protection and confidentiality of personal data should be a top priority for all types of organisations, the group agreed. — General public, Citizens’ Council, NICE, UK, 2015. (31, p.27) |
| That the organisations involved be seen to be highly trustworthy and competent (19, 37, 38) | The reasons the juries gave for why organizations should not have access to the data included several that were the opposite of the reasons for access, such as organizations that did not clearly indicate that the primary use of the data is for public benefit, who may use the data solely for private gain or commercial profit, or who did not have a trusted track record for protecting data. — General public, Citizens Jury, UK, 2015. (37, p.9) |
| That the data be anonymised or aggregate data (16, 18-21, 24-26, 30, 31, 33, 35, 38) | ‘With critical credentials removed I can see enormous beneﬁt where data is used to forecast services for health and far beyond. I want my data to be part of that, as long as it’s not recognisable as being mine’. — Participant, general public, workshops, New Zealand, 2017. (19, p.12) |
| That researchers be vetted (24, 25) | ‘Licensing and regulation --- if people are doing it for good then they should be happy for it to be regulated. Every organisation should be licensed and regulated and audited to ensure that there is a level of openness’. — Patients (rare conditions), Sheffield, UK. (25, p.62) |
| That researchers’ activity on the system be tracked (25, 38) | Participants pointed out that the individual themselves may not be trustworthy. Many are reassured by the suggestion that every time somebody has access they should be required to enter a log. The public like that if anything goes wrong it is easy to trace the user and that in turn acts as a driver of good practice and means it is possible to monitor and enact consequences for excessive use. — Public and patients, workshops, UK, 2016. (25) |
| That the data analysis be undertaken in the public or university sector (24, 25) | ‘The pharma company should pay for it, the regulator or academics should do it’. — General Public, Sutton Coldfield, UK, 2016. (25, p.51) |
| Scientific Integrity |  |
| That any research using data linkage and sharing be scientifically robust (19, 27, 31) | Good scientific practice to ensure the accuracy and validity of research design and data analysis. The Council felt that research that does not produce any useful findings because it is not scientifically robust is a waste of time and resources. — General public, Citizens’ Council, NICE, UK, 2015. (31, p.43) |
| Accountability and Oversight | |
| That there be independent oversight or a strong regulatory body overseeing the research (16, 24, 25, 30, 33, 35, 37, 38) | ‘I found it encouraging that the information and privacy commissioner has an oversight over it and it renews every 3 years. I found that encouraging. Someone’s keeping an eye on it’.— Participant, general public, group 1, Focus groups, Sudbury, Canada, 2017. (18, p.E43) |
| Criminal penalties (fines, prison sentences) in the event of wrongdoing (24, 25, 30) | Do you think that perhaps the reason we’re not happy with many people having that level of power over our data, partly because we don’t believe that the penalties for misusing data are severe enough? I mean, for me, that’s a crucial point, I actually think there would be less mismanagement of data if the penalties of knowingly selling or giving away personalised information carry far greater criminal penalties [.. . Currently] They don’t, I mean, you’re not going to go to jail for it! Whereas, perhaps if you did people would be less likely to, you know, purposely sell personalised information. — Male1, social service researcher, Scotland, 2016. (30) |
| That collection of and access to data be appropriate – sufficient for purpose and no more (29, 31, 37) | Typically, these organizations clearly demonstrated that the primary goal for using the data was for public benefit (such as improved medical care and treatments, improved public health, or management of public funds) and made a clear and compelling case for why they need these patient records. They provided clear justification for how and why the data would be used, why it was relevant to their efforts, with whom it will be shared, and only access records they needed to perform their data analysis and could not get adequate data from other sources. — General public, Citizens’ Juries, UK, 2018. (37, p.9) |
