## Supplementary material for "“Public attitudes to sharing government data with private industry: a systematic scoping review”": Cover letter

30^th^ May 2022

Dear Sir/ Madam

**Re Submission of Manuscript**

### Thank you for the opportunity to submit our manuscript ‘Public attitudes to sharing government data with private industry: A scoping review’

to your journal for consideration. We look forward to your feedback.

Yours sincerely

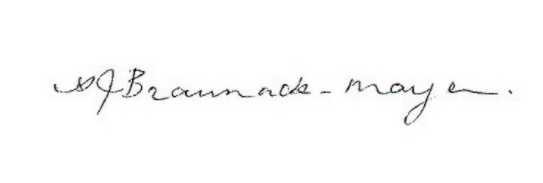

Prof Annette Braunack-Mayer
