## Supplementary material for "“Public attitudes to sharing government data with private industry: a systematic scoping review”": Table 1 Inclusion _Exclusion Criteria

Table 1: Inclusion and exclusion criteria for selection of articles

|  | Inclusion Criteria | Exclusion Criteria |
| --- | --- | --- |
| Publication Date | March 2014 – March 2019 | Before March 2014 |
| Document Type | Journal articles, conference paper, review, book chapter, book, article in press and reports (govt. and non-govt.) | Thesis, blog, PowerPoint slide, and research agencies websites |
| Study Design | Empirical | Conceptual |
| Study Population | Community members, public, patient groups | Clinical stakeholders, private industry |
| Research Topic | Articles were selected because, with respect to the use of any administrative data* by private industry** for therapeutic development***, they described empirical examples or understandings of:   - community attitudes - public interest or public benefit - social licence   Private uses of public data for therapeutic development  Private uses of public data outside therapeutic development in the biomedical/health sector  Studies that were unclear about whether data were actually held in the public sector, but where this could be surmised (such as in the case of a cancer registry)  US studies where it was not clear if the data being shared (electronic health record data or routinely-collected administrative data) were held in the public sector or by a private managed-care consortium.  Studies involving expert or stakeholder opinions if public and expert/stakeholder responses were reported separately. | Outside of therapeutic development: The paper is not related to data sharing in the health sector i.e. it is about data sharing in an alternate sector e.g. food, financial or environmental sectors or for insurance or marketing within or outside the health sphere.  Technical ONLY: The paper ONLY describes technical methods for analysing, sharing and linking data  Data sharing at individual level – e.g. sharing information between patients and doctors |

*Administrative health data: information collected and held in the public sector including, but not restricted to electronic health care records (including general practice, hospitals and allied health), registries and national disease databases.

**Private industry: companies developing, producing and selling pharmaceutical and medical devices (health insurance and marketing companies were excluded).

***Therapeutic development particularly focused on the development of medicines and medical devices
