## Supplementary material for "“Public attitudes to sharing government data with private industry: a systematic scoping review”": PRISMA Flow Chart

Cinahl: 365

Scopus: 1698

PubMed: 726

Total Relevant: 13

WOS: 1,745

Google Scholar: 992

Google Advanced: 170

Total: 5,526

Duplicate: 1,352

Total: 4,200

Total Relevant (abstract and heading screened): 81

Cinahl: 24

Scopus: 70

PubMed: 71

Google Scholar: 998

Total: 1,262

Duplicate: 108

Total: 1,153

Total Relevant (abstract and heading screened): 107

WOS: 99

Google Advanced: 123

Total Relevant: 27

Total: 228

Duplicates: 11

Total for full text reading: 217

Pearled: 4

**Total to be included: 33**

**Figure 1: PRISMA**

Search 1: Community Attitudes

Search 2: Public Interest & Social Licence
