## Supplementary material for "“Public attitudes to sharing government data with private industry: a systematic scoping review”": Table 3 Included articles

| Title of article (grouped where relevant into studies) | Reference | Date of data collection | Country | Quality score | Publics engaged | Method of data collection | Sample number | Recruitment strategy |
| --- | --- | --- | --- | --- | --- | --- | --- | --- |
| Moving from trust to trustworthiness: Experiences of public engagement in the Scottish health informatics programme | (Aitken et al., 2016) | 2010 -2011 | UK | 7 | Public, patients (including some researchers) | Focus groups | 50 | Diverse range of backgrounds recruited through existing groups |
| Public preferences regarding data linkage for health research: a discrete choice experiment | (Aitken, McAteer, et al., 2018) | 2016 | UK | 7 | public | Online survey/discrete choice experiment | 1004 | Online panel |
| Who benefits and how? Public expectations of public benefits from data-intensive health research | (Aitken, Porteous, et al., 2018) | 2017 | UK | 7 | public | Deliberative workshops | 69 | Quota-based representative sample of population |
| Challenges arising when seeking broad consent for health research data sharing: a qualitative study of perspectives in Thailand | (Cheah et al., 2018) | 2017 | Thailand | 7 | clinical trial participants incl. healthy volunteers, community members with research interest | Semi-structured interviews  Focus groups | 18  19 | Through existing research networks |
| Patient/family views on data sharing in rare diseases: Study in European LeukoTreat project | (Darquy et al., 2016) | 2012 | Europe | 6 | patients and their families | Online survey | 195 | Via referrals & conference |
| A path to social licence &  Our data, our way | (Brown et al., 2017; Data Futures Partnership, 2017) | 2017 | NZ | 7 | public including dedicated engagement with Maori | Workshops  Online survey  Hui  Online hui  Online survey | 379  4033  94  60  533 | Through community groups |
| Public attitudes to data integration: Report prepared for Statistics New Zealand | (Davison et al., 2015) | NG | NZ | 7 | public | Narrative interviews Workshops | 29  30 | Through community groups – active targeting of Māori, Pasifika, unemployed, self-employed, retired |
| De-identified genomic data sharing: The research participant perspective | (Goodman et al., 2017) | 2013 | USA | 6 | public, patients and their families | Online survey | 450 | Cancer registry participants and controls |
| The importance of purpose: moving beyond consent in the societal use of personal health data | (Grande et al., 2014) | 2012 | USA | 7 | public | Online survey | 3,064 | Nationally representative sample - random digit dialling & address sampling |
| Wellcome Trust Monitor Report Wave 3: Tracking public views on science and biomedical research | (Huskison et al., 2016.) | 2016 | UK | 7 | public | Online survey | 1,524 | Nationally representative sampling - address sampling |
| The One-Way Mirror - Public attitudes to commercial access to health data | (Ipsos MORI, 2016) | 2015 | UK | 6 | public, patients | Deliberative workshops and workshops (incl. clinicians)    Face to face survey | 246  2,017 | Quotas to provide diversity with on-street recruitment |
| Research participants’ attitudes towards the confidentiality of genomic sequence information | (Jamal et al., 2014) | 2011-2012 | USA | 7 | public, patients | Semi structured phone interview | 30 | Enrolees in genomic research project |
| Older adults willingness to share their personal and health information when adopting healthcare technology and services | (Kim & Choi, 2019) | 2017 | South Korea | 3 | older adults | Face to face survey | 170 | Random on-street recruitment |
| Attitudes towards personal genomics and sharing of genetic data among older Swiss adults: A qualitative study | (Mählmann et al., 2018) | 2013-2014 | Switzerland | 6 | older adults | Face to face semi-structured interviews | 40 | Attendees at Seniorn universitat |
| You should at least ask'. The expectations, hopes and fears of rare disease patients on large-scale data and biomaterial sharing for genomics research | (McCormack et al., 2016) | 2016(2014) | International | 6 | patients | Focus groups | 52 | Attendees at EURORDIS meeting and summer school |
| Canadians' views about using big data in health research from a national online survey: A partnership of patient consumers and researchers | (McCormick et al., 2018) | 2017 | Canada | 5 | public, patients | Online survey | 151 | Targeted websites, email lists, social media |
| Stakeholders’ views on data sharing in multicenter studies | (Mazor et al., 2017) | 2015 | USA | 7 | patients | Face to face or telephone semi-structured interviews | 15 | Through engaged patient groups |
| Sharing of Big Data in Healthcare: Public opinion, trust and privacy considerations for health informatics researchers | (Moss et al., 2017) | NG | UK | 3 | public | Face to face survey | 37 | Through science festival |
| Attitudes towards data collection, ownership and sharing among patients with Parkinson's disease | (Mursaleen et al., 2017) | 2016 | International | 4 | patients | Online survey | 310 | Charities and support groups |
| What ethical and practical issues need to be considered in the use of anonymised information derived from personal care records as part of the evaluation of treatments an delivery of care | (NICE Citizens Council, 2015) | 2015 | UK | 7 | public | Deliberative meeting | 30 | Standing citizens’ council |
| Patient and public views about the security and privacy of Electronic Health records in the UK: results from mixed methods study | (Papoutsi et al., 2015) | 2011-2013 | UK | 4 | public, patients | Cross-sectional survey  Focus groups | 2761  114 | Through GP surgeries and hospital waiting areas |
| Social licence and the general public's attitudes toward research based on linked administrative health data: a qualitative study | (Paprica et al., 2019) | 2015 & 2017 | Canada | 7 | public | Focus group | 65 | Purposive sampling for diversity – age, education, income, levels of trust |
| Big desire to share big health data: A shift in consumer attitudes towards personal health information | (Pickard & Swan, 2014) | 2014 | International | 4 | public | Online survey | 128 | Crowdsourced through online group |
| Public preferences regarding informed consent models for participation in population-based genomic research | (Platt et al., 2014) | 2007-2008 | USA | 7 | public | Survey | 3347 | Nationally representative sample - random digit dialling |
| Public trust in health information sharing: A measure of system trust | (Platt et al., 2018) | 2014 | USA | 6 | public | Online survey | 1011 | Nationally representative sample - email |
| Public attitudes to data sharing in northern Ireland &  Public attitudes to data sharing in Northern Ireland: Findings from the 2015 Northern Ireland Life and Times Survey | (Robinson & Dolk, 2015; Robinson et al., 2018) | 2015 | UK | 6 | public | Survey | 1202 | Systematic random sampling of addresses |
| Mental health service users' perceptions of data sharing and data protection: a short qualitative report | (Satinsky et al., 2018) | 2017 | UK | 1 | consumers | Focus group | 8 | Social media |
| Patient perspectives on sharing anonymized personal health data using a digital system for dynamic consent and research feedback: a qualitative study | (Spencer et al., 2016) | 2016 | UK | 7 | patients | In-depth interviews  Focus groups | 26  14 | Teaching hospital outpatient clinic & patient public involvement health network |
| “You hoped we would sleep walk into accepting the collection of our data”: Controversies surrounding the UK care.data scheme and their wider relevance for biomedical research | (Sterckx et al., 2016) | 2013-2015 | UK | 7 | public | Qualitative analysis of blogs | 256 | Blogs |
| Investigating the extent to which patients should control access to patient records for research: A deliberative process using citizen juries | (Tully et al., 2018) | NG | UK | 7 | public | Citizen juries | 34 | Reflecting UK national demographic profile |
| Data Sharing and Technology: Exploring the attitudes of people with asthma | (West & Cumella, 2018) | 2018 | UK | 5 | patients | Online survey | 3054 | Annual survey |
| ODI Survey reveals British consumer attitudes to sharing personal data | (United Kingdom Government, 2018) | 2017 | UK | 6 | public | Online survey | 2023 | Members of YouGov Plc UK panel |
| The use of information for diabetes research and care: Patient views in West London | (Zalin et al., 2016) | 2011-2012 | UK | 4 | patients | Cross sectional survey  Focus group | 404  6 | Hospitals and GP surgeries |
