## Supplementary material for "“Public attitudes to sharing government data with private industry: a systematic scoping review”": Table 4 Participants willing to share

**Table 4:** Proportion of participants willing to share administrative health data with specific groups (all quantitative studies asking this question)

|  | Family | Own health provider | Health services | Government departments | Academic researchers/ universities | Government researchers | Private industry | Commercial organisations such as insurance companies | Charities/ non-profit organisations |
| --- | --- | --- | --- | --- | --- | --- | --- | --- | --- |
| Northern Ireland adults n=1202 (21)(21) | - | 91% | 86% | 73% | 72% | - | 75%* | 41% | 51% |
| Ipsos MORI for Wellcome Trust n= 2017 (4)(4) | - | - | - | - | - | 54% | 53% | 26% (insurance)  37% (marketing) | - |
| USA Crowd sourced n=128 (44)(44) |  | 84% | - | 28% | 79% |  | 24% | 14% | - |
| USA genetics registry participants  n=450 (30)(30) | - | - | - | - | 90-93 (same university) 76-87% (other universities) | 64-76% | 16-24% | - | 84-91% |
| European leukodystrophy patients/carers n=195 (28)(28) | 37% | 91-96% (specialist)  72-76% (family doctor) | - | - | 64-90% | - | 61-65% | - | - |
| Maori, New Zealand n=533 (20)(20) | 82% | - | - | 77% | - | - | 46% | - | - |
| South Korea, older adults, n=170 (34)(34) | 80%* | - | 66%* | - | 39%* | 24%* | 25%* | 19%* | - |
| London patients with diabetes n=404 (52)(52) | - | - | 51% (anonymised)  29% (identifiable) | - | - | - | 45% (anonymised)  15% (identifiable) | - | 49% (anonymised)  17% (identifiable) |

*Question relates to a scenario involving use of deidentified data where company is working for a cure for Alzheimer’s Disease
